# Explainable, Lightweight Deep Learning for Colorectal Cancer Microsatellite Instability Screening in Low-Resource Settings

**DOI:** 10.64898/2026.04.18.26350809

**Authors:** Olubola Titilope Adegbosin, Hiran Patel

**Affiliations:** School of Mathematics and Computer Science, Faculty of Science and Engineering, University of Wolverhampton, Wolverhampton, West Midlands, WV1 1LY, United Kingdom; Department of Radiation/Clinical Oncology, University of Benin Teaching Hospital, Benin City, Edo State, Nigeria

**Keywords:** artificial intelligence, colorectal cancer, deep learning, digital pathology, explainable AI, Grad-CAM, histopathology, medical AI, microsatellite instability, resource-limited settings

## Abstract

**Background:** Microsatellite stability status determination is important for prognostication and therapeutic decision making in colorectal cancer management, but the conventional methods for this assessment are not readily available, especially in low- and middle-income countries. Deep learning (DL) models have been proposed for addressing this problem; however, potential computational cost due to model complexity and inadequate explainability may limit their adoption in low-resource settings. This study explored the potential of explainable lightweight models for detection of microsatellite instability in colorectal cancer.

**Methods:** DL models were trained using a public dataset of colorectal cancer histology images and then used to classify a set of test images into one of two classes: microsatellite instability or microsatellite stability. The models were compared for efficiency. Gradient-weighted class activation mapping (Grad-CAM) was used to interpret the models’ decision making.

**Results:** The simpler convolutional neural network (CNN) trained from scratch had modest performance (accuracy=0.757, area under receiver-operating characteristic curve [AUROC]=0.840). With an attention mechanism added, these values increased, but specificity and sensitivity reduced. Pretrained models performed better than the ones trained from scratch, and EfficientNet_B0 had the best balance of high performance and low computational requirements (accuracy=0.936, AUROC=0.990, negative predictive value=0.923, specificity=0.953, 4,010,000 trainable parameters, 0.38 gigaFLOPs). However, a simple CNN model with attention mechanism had the best interpretability based on Grad-CAM.

**Conclusion:** This study demonstrated that DL models that are lightweight when compared to previously proposed ones can be useful for colorectal cancer microsatellite instability screening in resource-limited settings while balancing performance and computational efficiency.

## 1. INTRODUCTION

Colorectal cancer is the third-commonest cancer and second-leading cause of cancer-related mortality globally [1], and its incidence and mortality rates are projected to increase by 63% and 73% respectively in the next two decades [1].

Colorectal carcinogenesis is an interplay of multiple genetic abnormalities, some of which have been studied as therapeutic targets. Among these are mismatch repair (MMR) genes, which are responsible for maintaining the DNA structure and preventing microsatellite instability (MSI) by correcting errors in non-coding DNA regions called microsatellites; however, MMR genes themselves may be mutated, resulting in the MSI that underlies oncogenesis in some cancers [2, 3]. MSI is most commonly found in colorectal cancer, being present in about 15% of cases globally [4]. The burden is even higher in some populations: for instance, two studies from Nigeria report prevalences of 28.1% and 43% [5,6]. Colorectal cancer with MSI has favourable prognosis in the early stage and is amenable to immunotherapy [4,7,8], while colorectal cancer with microsatellite stability (MSS) does not benefit from this expensive treatment. Moreover, MSI in colorectal cancer may be an indication of the familial Lynch syndrome [3]. Hence, the microsatellite stability status of colorectal cancer is of therapeutic and prognostic importance.

Conventionally, MSI status is assessed via two methods [7]. The direct method, which is also considered the gold standard, involves polymerase chain reaction to determine the structures of MMR genes. This method requires expertise and equipment that are not readily available, especially in low-resource settings. The indirect method involves the relatively cheaper immunohistochemistry, and even this is not readily accessible or affordable for patients in climes where healthcare financing is grossly inadequate. Next-generation sequencing can also be used to detect MSI [9], but this is even more high-end than the two methods described previously. This poor access to MSI diagnostic resources in low-resource areas coincides with the higher disease burden in the same settings, and it contributes to poor patient outcomes due to delayed or inadequately informed clinical decisions. Therefore, strategies to facilitate access to MSI detection assays are particularly needed in low-resource settings.

The past few decades have been characterized by the application of artificial intelligence (AI) in medicine. Particularly, with respect to colorectal cancer, AI has been used extensively for various purposes [10], including to aid screening [11], predict risk [12], diagnose [13], and predict therapeutic response and survival [14]. There is a body of existing work on the use of deep learning (an AI subspecialty) to assess microsatellite stability status in colorectal cancer, using haematoxylin-eosin (H&E)-stained histology images [15-20]. With respect to real-world applications, Owkin Inc. [21] developed MSIntuit, a deep learning tool for screening for MSI in colorectal cancer, and this tool has been clinically validated [22]. The results obtained from these studies have been remarkable, with accuracies as high as 90%, but the studies employed very deep neural networks, which translates to high computational costs that may be restrictive in low-resource settings. Most studies also did not attempt interpretability, which is an important determinant of adoption in places where AI literacy and interest are low.

The current study aims to i) train lightweight deep learning models for determination of microsatellite stability status of colorectal cancer, ii) compare the lightweight models to deeper ones in terms of performance metrics and computational efficiency, and iii) explore explainability of the models’ decisions.

## 2. METHODS

### 2.1. Experimental design

In this study, we approached MSI detection as a binary classification task. We trained deep learning models to receive H&E-stained histology images as input and classify them into one of two classes: MSI or MSS. The models were trained with images labelled as MSI or MSS and then validated on another set of images. Model performance was evaluated with a third set of images (test dataset) that the models were not exposed to during training or validation. The flowchart of the experimental design is shown in Fig.1.

**Fig. 1.**
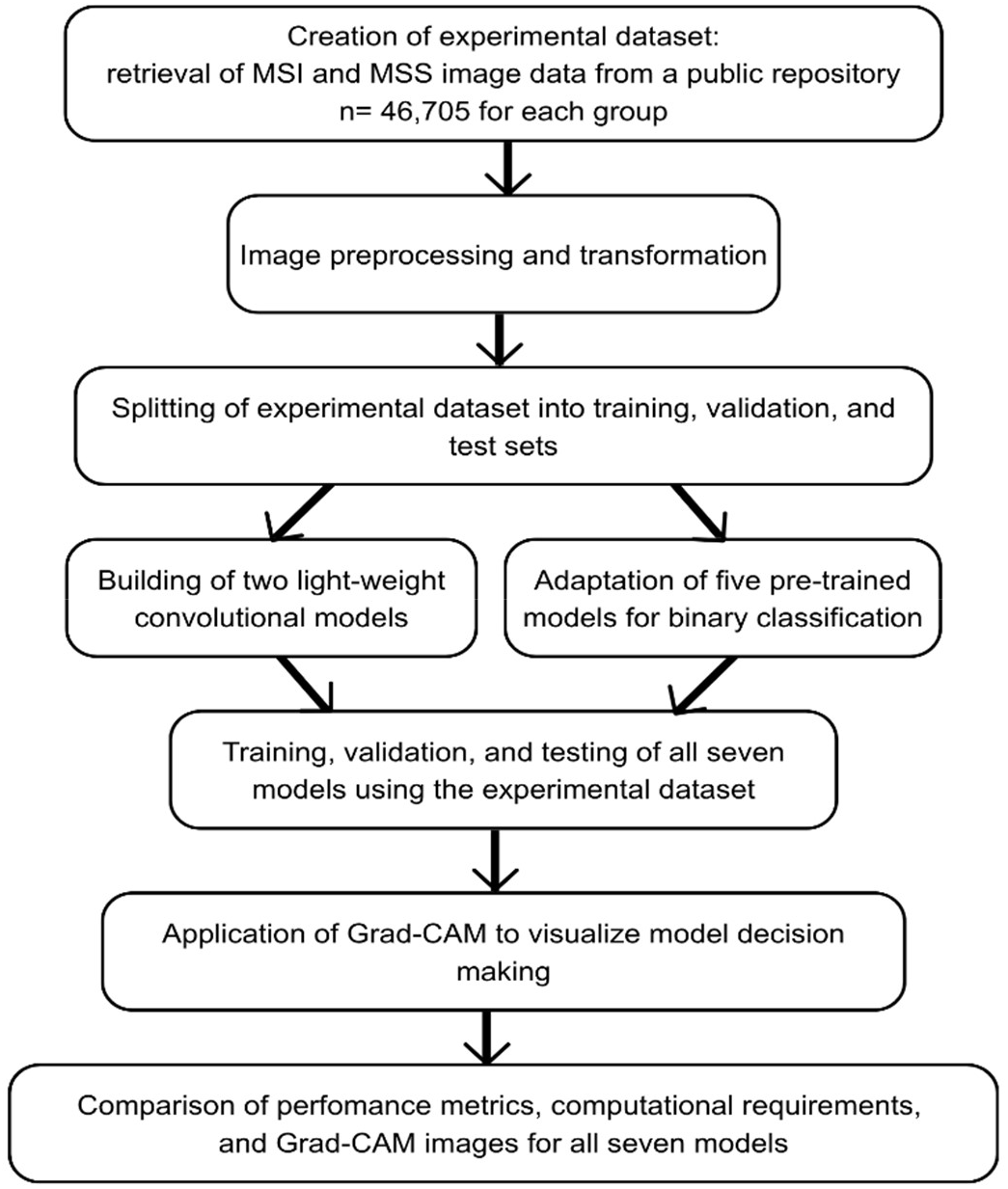
Flowchart of the experimental design of this study. MSS: microsatellite stability; MSI: microsatellite instability; Grad-CAM: gradient-weighted class activation mapping

### 2.2. Data collection and preprocessing

The data used was obtained from a publicly available repository [23] that was derived from de-identified histological images of colorectal cancer specimens in The Cancer Genome Atlas (TCGA). The whole dataset contained 411,890 unique, 3-channel image patches derived from whole-slide images. The images had been resized to 224 pixels x 224 pixels at a resolution of 0.5 µm/pixel and colour-normalized using the Macenko method. For this study, only a portion of the dataset was used (46,705 images each of MSS and MSI colorectal cancer). An 80:10:10 split was applied to the dataset to generate training, validation, and test datasets, respectively. The images were augmented by applying horizontal flip, brightness, and contrast transformations, after which they were converted to tensors and then normalized.

### 2.3. Model architecture

Two models trained from scratch and five pretrained models were explored. The first from-scratch model is a simple convolutional neural network (CNN). It consists of three convolutional layers (with kernel size of 3×3 and padding of 1), each followed by batch normalization to stabilize learning, rectified linear unit activation to prevent overfitting and vanishing gradients, and max pooling (a downsampling operation with kernel size of 2 and stride of 2 that helps the model to focus on the strongest activations in each layer). Early layers capture simple patterns, while deeper layers identify more complex structures, such as tissue architecture or tumour characteristics. After the convolutional blocks, the output is flattened into a 1D vector and passed through two dense layers: the first layer has 512 neurons, and the second produces a single output. This structure allows the model to learn hierarchical features from the input image; these are then transformed through the dense layers to produce a final scalar value for binary classification.

The CNNAttentionOnly model built on Simple_CNN by adding a self-attention mechanism to enhance feature learning. This mechanism allowed the model to consider relationships between different regions of the image, so that each part can be influenced by spatial patterns across tumour sections. The attention output was then integrated into the CNN’s dense layers to generate the final prediction. Notably, the residual connection between the attention input and output (which is usual in this kind of model) was omitted—only the scaled attention output was passed to the dense layers of the CNN, as our preliminary experiments showed that relying solely on the attention-enhanced features improved model performance. The architecture of the CNNAttentionOnly model is shown in Fig. 2.

**Fig. 2.**
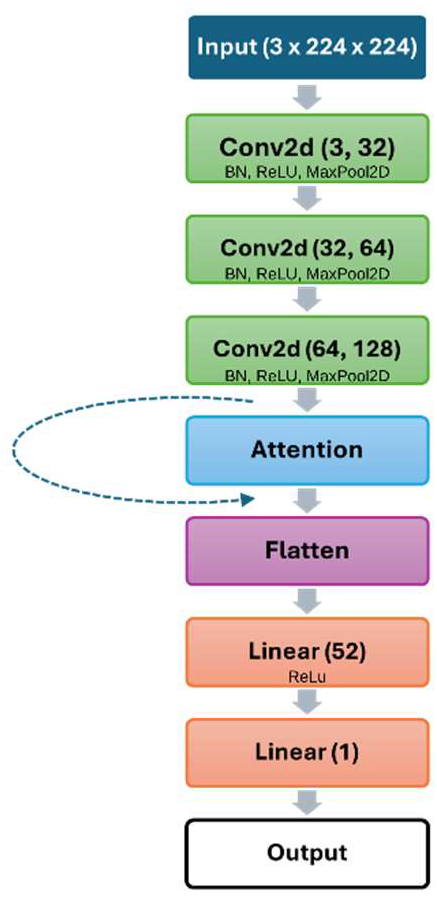
Architecture of the CNNAttentionOnly model used in this study. For each convolutional layer, kernel_size=3 and padding=1. The curved arrow with broken lines represents the usual residual connection between attention input and output, which was omitted in this architecture. This architecture without the attention block is that of the simple_CNN model. BN: batch normalization; ReLU: rectified linear unit activation; MaxPool2D: 2D max pooling

Five pre-trained models were also deployed, namely VGG16, VGG19, ResNet18, ResNet50, and EfficientNet_B0. VGG16 is a deep CNN introduced by the Visual Geometry Group at the University of Oxford in 2014 [24] to explore how network depth affects performance in image analysis. It has a straightforward, uniform structure, and its deep architecture allows high performance, though it is computationally expensive compared to recent models. VGG19 is its 19-layer variant. ResNet-18 and ResNet-50 are the 18- and 50-layer members of the ResNet (Residual Network) models, which were introduced in 2016 [25]. They include residual connections, which help the model learn complex patterns without losing important information. EfficientNet-B0 is the simplest model in a family of highly efficient CNNs developed by Google AI [26]. It was designed to deliver high accuracy while requiring less computational power, making it practical for use in resource-limited environments. Its architecture allows it to capture rich image features without unnecessary complexity. The final layer of each pretrained model was modified for binary classification.

For training the models, Adam optimizer was used, and heuristic search was performed to determine optimal model hyperparameters (learning rate and weight decay). For the from-scratch models, learning rate and weight decay of 1e-4 were optimal, while for the pre-trained models, 1e-5 and 1e-4 were the optimal learning rate and weight decay, respectively. The loss function used was binary cross entropy with logits. Training batch size was 32. Early stopping was implemented to interrupt the training loop if the validation accuracy did not improve after 3 consecutive epochs.

### 2.4. Model evaluation

Model performances were evaluated using accuracy, area under receiver operating characteristic curve (AUROC), negative predictive value (NPV; chosen because it translates to minimal false negatives), sensitivity, and specificity. The computational requirements of the models, in terms of training duration, number of trainable parameters, and number of floating point operations (FLOPs), were recorded. FLOPs refer to the total number of operations (such as addition, multiplication, and division) that a model performs during one training step: more FLOPs generally mean that the model is more complex and computationally expensive [27].

### 2.5. Model interpretation

Gradient-weighted class activation mapping (Grad-CAM) was used to overlay heatmaps on images to show which regions contributed most to each model’s prediction, thus allowing visual linking of model decisions to relevant tissue features [28].

### 2.6. Computational environment

All models were built and/or trained using PyTorch 2.6.0+cu126. Performance metrices were calculated using parameters from sklearn.metrics. The codebase was implemented in Python 3.11.9, using a Jupyter Notebook environment on a local workstation equipped with an NVIDIA GeForce RTX 4060 GPU and CUDA 12.8.

### 2.7. Ethical considerations

The dataset used in this study was derived from the TCGA datasets, and no attempts were made to re-identify any individual. The original TCGA participants provided broad consent for the research use of their biospecimens and data, permitting secondary analyses such as those presented here.

## 3. RESULTS

### 3.1. Performance metrics

Simple_CNN had an accuracy of 0.757, AUROC of 0.840, and NPV of 0.747. CNNAttentionOnly had higher accuracy and AUROC but lower NPV, specificity, and sensitivity than Simple_CNN. Overall, the pretrained models (VGGs, EfficientNet, and ResNets) performed better than the from-scratch models, with accuracy >90%, NPV ranging from 0.884 to 0.952, and AUROC of at least 0.98. ResNet18 had the second-highest specificity (0.952) after EfficientNet_B0 (0.953), but its other metrices were lower than those of the other models. EfficientNet_B0 had lower NPV and sensitivity that most of the other models; for the other metrices, it had the highest values. Details of model performance are given in Table 1. The confusion matrices for the models are shown in Fig. 3.

**Table 1.** Performance metrics of the models used in this study.

| **Model** | **Accuracy** | **NPV** | **AUROC** | **Sensitivity** | **Specificity** |
| --- | --- | --- | --- | --- | --- |
| *From Scratch* |  |  |  |  |  |
| Simple_CNN | 0.757 | 0.747 | 0.840 | 0.736 | 0.778 |
| CNNAttentionOnly | 0.832 | 0.733 | 0.930 | 0.725 | 0.753 |
| *Pretrained* |  |  |  |  |  |
| ResNet18 | 0.914 | 0.884 | 0.980 | 0.875 | 0.952 |
| ResNet50 | 0.934 | 0.935 | 0.990 | 0.935 | 0.933 |
| VGG16 | 0.934 | 0.930 | 0.980 | 0.929 | 0.938 |
| VGG19 | 0.935 | 0.952 | 0.990 | 0.953 | 0.917 |
| EfficientNet_B0 | 0.936 | 0.923 | 0.990 | 0.919 | 0.953 |

AUROC: area under the receiver operating characteristic curve; CNN: convolutional neural network; NPV: negative predictive value

**Fig. 3.**
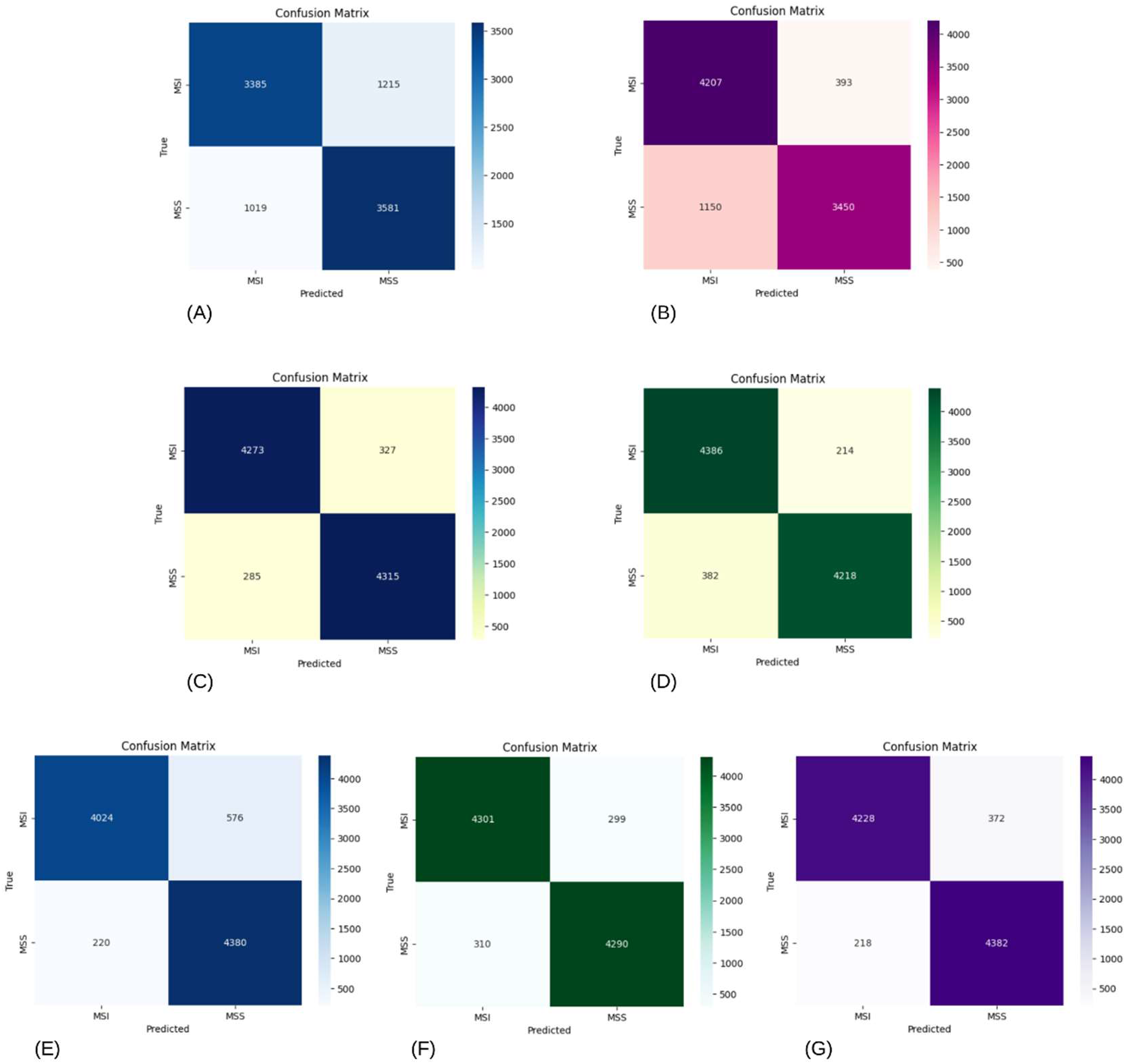
Confusion matrices of the models tested. (A) Simple_CNN, (B) CNNAttentionOnly, (C) VGG16, (D) VGG19, (E) ResNet18, (F), ResNet50, and (G) EfficientNet_B0 MSI: microsatellite instability; MSS: microsatellite stability

### 3.2. Computational requirements

The models were also compared for efficiency. VGG19 had the longest training time (163 minutes) and largest number of trainable parameters and FLOPs. Simple_CNN and ResNet18 had the shortest training time (64 minutes), while EfficientNet_B0 had the smallest number of trainable parameters and FLOPs (Table 2).

**Table 2.** Computational requirements of the deep learning models used in this study.

| Model | Training Time<br>(minutes) | Number of Trainable<br>Parameters (million) | Giga FLOPs |
| --- | --- | --- | --- |
| *From Scratch* |  |  |  |
| Simple_CNN | 64 | 51.47 | 0.57 |
| CNNAttentionOnly | 134 | 51.50 | 0.58 |
| *Pretrained* |  |  |  |
| ResNet18 | 64 | 11.18 | 1.82 |
| ResNet50 | 122 | 23.51 | 4.13 |
| VGG16 | 148 | 134.26 | 15.47 |
| VGG19 | 163 | 139.57 | 19.63 |
| EfficientNet_B0 | 89 | 4.01 | 0.38 |

CNN: convolutional neural network; FLOPs: floating point operations

### 3.3. Interpretability

The heatmap for CNNAttentionOnly showed that widespread areas that contained nucleated cells contributed the most to the model’s decision making, while that of Simple_CNN showed a reverse pattern. Heatmaps for the ResNet models showed the most activity in focal areas of nucleated cells; VGG heatmaps showed focal areas of maximum activity along the boundaries of nucleated cellular regions and vacant areas of the images. EfficientNet_B0 heatmap showed maximum activity in focal areas also, including areas that are not populated by (nucleated) cells (Fig. 4). For comparison, images from immunohistochemistry for microsatellite status of colorectal cancer [29] have been provided.

**Fig. 4.**
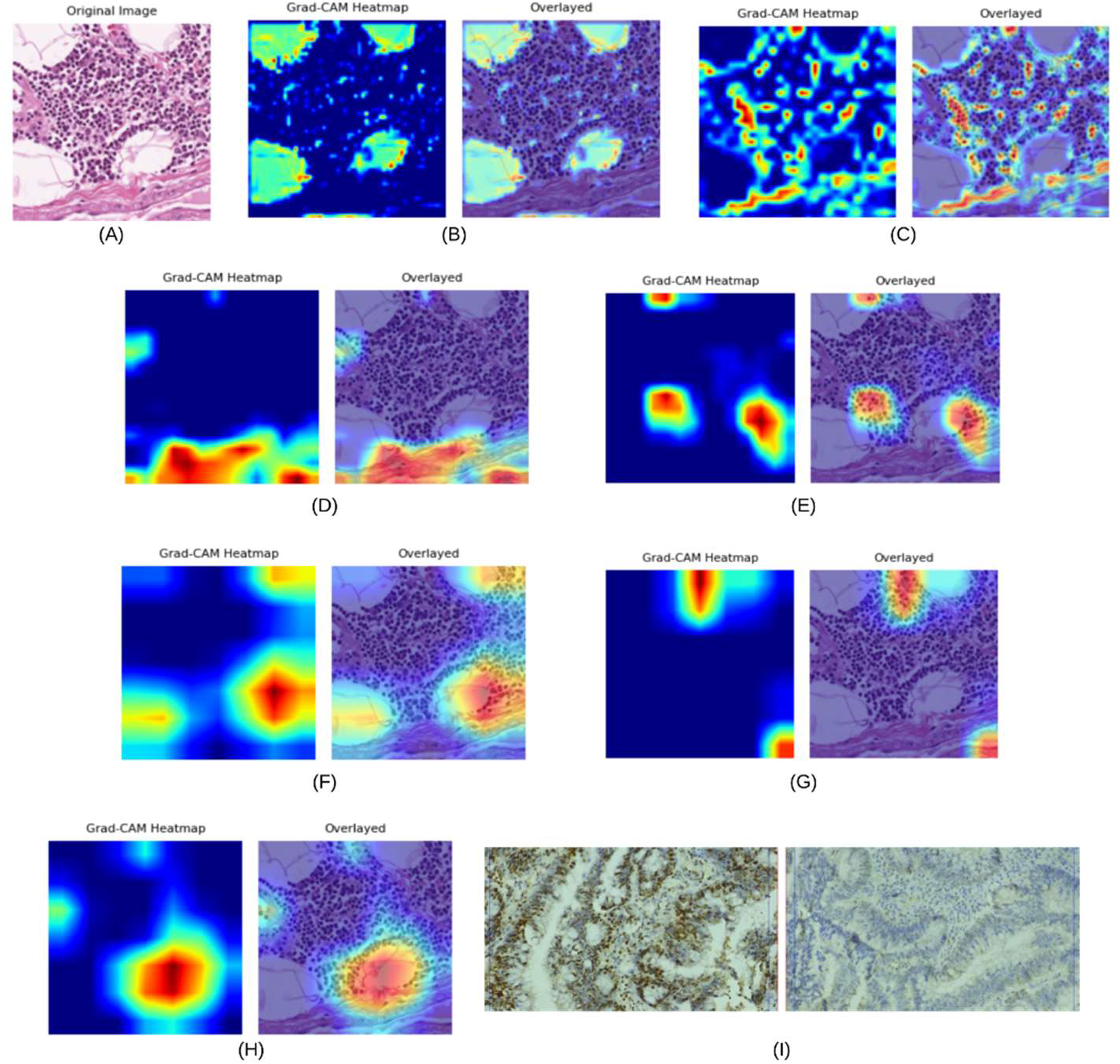
Visualization of decision making by the models using gradient-weighted class activation heatmaps. (A) Histology image of colorectal cancer with microsatellite instability. Gradient-weighted class activation heatmaps and corresponding image plus heatmap overlay for (B) Simple_CNN, (C) CNNAttentionOnly, (D) VGG16, (E) VGG19, (F) ResNet18, (G), ResNet50, and (H) EfficientNet_B0. Warm colours depict areas that contribute the most to model decision making. For comparison, (I) cropped immunohistochemistry image from Yassen et al. (https://creativecommons.org/licenses/by/4.0/) [29] showing intact nuclear expression (brown colour) of a mismatch repair protein (suggesting microsatellite stability) in the left panel versus loss of nuclear expression of the mismatch repair protein (suggesting microsatellite instability) in the right panel.

## 4. DISCUSSION

This study aimed to explore and evaluate computationally inexpensive deep learning models for MSI screening in colorectal cancer. Our findings showed that EfficientNet_B0 was the most efficient of all seven models tested, balancing high performance metrics with low computational requirements.

The performance metrics obtained in the current study are comparable to what have been reported by other researchers. Simple_CNN performed as well, in terms of AUROC, as more complex models described by some previous researchers on this topic [17,18] who reported AUROC ranging from 0.84 to 0.88; however, its performance is inferior to those of the MSINet proposed by Yamashita et al. (AUROC = 0.931) [15] and modified Shufflenet architecture proposed by Echle et al. (AUROC = 0.94) [16]. With the added attention mechanism, the performance metrics improved: CNNAttentionOnly correctly classified about 80% of the data, and the AUROC of 0.930 shows a solid ability to differentiate between classes; however, it had lower specificity, sensitivity, and NPV than Simple_CNN. A possible reason for this observation is that attention mechanisms may amplify noise or irrelevant features in the training dataset, leading to increased false positives and false negatives (evident when the confusion matrices of Simple_CNN and CNNAttentionOnly are compared) that manifest as reduced specificity and sensitivity [30, 31]. Another potential explanation is that attention mechanisms are sensitive to feature alignment across spatial contexts, and this can lead to less effective classification if the training data exhibit significant histomorphologic variability [32]. It is important to recall here, that as stated in the Methods section, omission of the residual connection between the input and output of the attention mechanism of the CNNAttentionOnly model resulted in better performance. This, as well as other studies [33], corroborates the inference that while attention mechanisms generally improve model performance, they may need to be adapted differently for different models and tasks, for optimal results.

The deeper pre-trained models tested had even better performances across all metrices, and they all had AUROC of at least 0.98. However, near-perfect AUROC does not translate directly to near-perfect performance [34], as is evident in the other performance metrices. In a clinical validation study, MSIntuit, which uses a multi-model network that includes a UNet, ResNet50, and multilayer perceptrons [22], had sensitivity of 0.98 and specificity of 0.46 [22]. The model that had the closest sensitivity to this in our study is VGG19 (0.953) and, surprisingly, all the models we tested had higher specificities (0.778-0.953) than reported for MSIntuit. There are several plausible explanations for this. First, simpler neural network architectures can be less prone to overfitting; hence, they may generalize better to intrinsic variations in the target data distribution by learning only the most salient features, rather than fitting noise or spurious correlations [35,36]. Second, the higher specificity of our models may be due to the tuning of MSIntuit to optimize sensitivity in the study referenced [22]. A standard 0.5 probability threshold was used for classification in our study, ensuring balanced decision-making at the pre-screening stage, so that MSI cases can be flagged for confirmatory testing. This threshold maintains clarity for clinical adoption. Nonetheless, rigorous external validation across varied clinical cohorts is required to ensure that the observed specificity advantage is reproducible and not attributable to sample bias or dataset artifacts [37].

The EfficientNet models are acclaimed for their good performance in computer vision tasks while using fewer parameters than older models like ResNet and VGG. On the ImageNet dataset on which both EfficientNet and ResNet were pretrained, although they achieved similar accuracies, EfficientNet_B0 used 20% of the parameters and 10% of the FLOPs used by ResNet-50 [26]. In this study, a similar trend was observed, with EfficientNet_B0 using the least number of parameters and FLOPs among all the models tested, while achieving the highest performance metrics (except for slightly lower NPV and sensitivity).

While the computational and infrastructural requirements of models described in previous related studies [15-19, 22] were not disclosed, high computational requirement is a reasonable assumption based on the models’ architectures [38], in contrast to the models tested in this study. The larger the number of trainable parameters of a model, the greater the need for high-performance graphics processing units or cloud-based infrastructure, which may not be readily available in low-resource settings [39]. With none of them exceeding 0.6 gigaFLOPs, the Simple_CNN, CNNAttentionOnly, and Efficientnet_B0 models can typically run in near real time on central processing units, consuming minimal electrical power and generating little heat [40]. Each would typically use less than one-third of the computational resources of the ResNet18 model, and this will allow deployment on modest hardware—an important determinant of adoption in low-resource settings.

Regarding model interpretability, CNNAttentionOnly had the most intuitive decision-making process as visualized with Grad-CAM heatmaps. During immunohistochemistry to determine the microsatellite stability status of colorectal cancer, pathologists evaluate the expression of MMR proteins (which are expressed on cell nuclei) in tumour tissue and compare it with expression in normal tissue, which serve as internal positive controls. Loss of staining in tumour cells for at least two MMR proteins, with preserved staining in normal cells, indicates MMR deficiency and suggests MSI, whereas intact staining in tumour cells indicates MMR proficiency and microsatellite stability [29]. Among the models compared, only the CNNAttentionOnly model showed a similar pattern of decision making, involving diffuse areas of nucleated cells; the more complex models showed localized heatmap patterns that do not correlate well with what is typically observed during immunohistochemistry. This tendency for simpler, from-scratch models to focus on visually prominent and humanly interpretable features, while complex pretrained models rely on abstract and generic features learned from large and diverse datasets, has been explained by other researchers [28]. The attention mechanism of CNNAttentionOnly may have also been responsible for it producing more humanly reasonable Grad-CAMs than the baseline CNN model.

Summarily, complex pretrained models generally performed better at classifying images as MSS or MSI in this study than the simpler models built from scratch, and EfficientNet_B0 is particularly desirable not only because of its excellent performance metrics, but also because it had the least computational requirements. However, the decisions of the pretrained models were not as intuitive as that of the CNN model with attention mechanism.

Summarily, this study has demonstrated that deep learning models that are lightweight when compared to previously proposed ones can be useful for colorectal cancer MSI screening while balancing performance and computational efficiency. This offers a potentially more affordable and accessible approach to MSI detection in colorectal cancer, which is especially relevant for settings where resource constraints limit widespread use of immunohistochemistry/PCR or high-end deep learning models/tools. Our study also showed that the best performing models are not necessarily the most explainable, whereas model explainability is key in critical applications such as that examined in this study.

### 4.1. Limitations

A key limitation of this study is that the models were trained on data that is not indigenous to the resource-limited populations among whom their use is being proposed, and this may limit generalizability. The TCGA dataset used is largely sourced from the United States of America, and majority of the samples are of Caucasian origin [41]. Moreover, the images used were not annotated to depict the various cell types that may be present in tumour sections, such as tumour, normal gland, or inflammatory cells. This information would have made discussions on model interpretability more robust. Future work directions would include replicating this study using indigenous datasets with cellular annotations to validate performance and improve evaluation of explainability. Further, it would be interesting to compare the decisions of the deep learning models with those of histopathologists.

Regardless of these limitations, our study puts forward a framework that can be improved upon to develop explainable and computationally inexpensive deep learning tools for MSI screening in colorectal cancer in low-resource settings. This could accelerate diagnosis, support timely clinical decision-making, and reduce both direct and indirect costs associated with colorectal cancer management.

## DECLARATIONS

### Authors’ contributions

Both authors conceptualized the study and designed the methodology. OTA implemented the code and wrote the manuscript draft. Both authors reviewed, edited, and approved the final manuscript draft.

### Data Availability

The dataset used in the study is available in the Zenodo repository, https://zenodo.org/records/2530835 [23].

